# Definitive Radiation Therapy With Dose Escalation Is Beneficial For Patients With Squamous Cell Cancer Of The Esophagus

**DOI:** 10.1101/2020.04.29.20077826

**Authors:** Sarbani Ghosh-Laskar, Naveen Mummudi, Saurabha Kumar, Mukesh Chandre, Shagun Mishra, Anil Tibdewal, JP Agarwal, Vijay Patil, Vanita Noronha, Kumar Prabash, Sabita Jiwnani, George Karimundackal, CS Pramesh, Prachi Patil

## Abstract

**OBJECTIVE:** We report the long term follow-up, toxicity, and outcomes of patients with localized squamous cell carcinoma of the esophagus (ESCC) who underwent definitive chemoradiotherapy (dCRT) at our institute.

**MATERIALS AND METHODS:** Patients diagnosed with carcinoma post cricoid, upper cervical and thoracic oesophagus and treated with dCRT treated between January 2000 and March 2012 were retrospectively analysed. Data was extracted from the hospital medical records and patient files. Patients deemed inoperable received upfront RT with or without concurrent chemotherapy and patients with borderline resectable and/or bulky disease received neoadjuvant chemotherapy followed by CRT or RT alone. Radiotherapy was delivered in two phases to a maximum dose of 63 Gy in daily fractions of 1.8 Gy using conventional or conformal techniques. Overall survival and progression free survival were defined from date of registration and were calculated by Kaplan-Meier method with comparisons between different subgroup performed using log rank test. All data were analyzed using SPSS Version 22.

**RESULTS:** Three hundred and fourteen patients with ESCC treated with dCRT were included in this analysis. Median age at presentation was 56 years and median Karnofsky Performance Status (KPS) at presentation was 70. Two-third of patients were treated with conformal technique alone or a combination of conventional and conformal technique. Median dose of radiation delivered was 60 Gy (range 30.6 Gy – 70 Gy). Neoadjuvant chemotherapy was administered in about 35% patients and 57% patients received concurrent chemotherapy. About 82% patients (77%) completed their planned treatment course; 10% patients required hospitalization during treatment due to complications and 7 patients did not complete treatment. Grade 1/2 dermatitis and mucositis was seen in 77% and 71% patients respectively. Grade 3 non-hematological and hematological toxicities were seen infrequently. Complete response at first follow up was observed in 56% of patients. At a median follow up of 56 months, 77 patients were alive with controlled disease. The 1-, 2- and 3-yr OS were 80%, 67% and 62% respectively. Median PFS was 28 months; 1-, 2- and 3-yr PFS were 66%, 52% and 46% respectively. A higher RT dose was found to be a significant predictor for OS and PFS on both uni- and multivariate analysis.

**CONCLUSION:** Our study highlights that delivery of higher RT doses (≥63□Gy) is feasible in this patient group and that a higher RT dose was associated with significantly better PFS and OS.

## INTRODUCTION

Esophageal cancer (EC) is the 12^th^ commonest cancer worldwide with about 600,000 new patients diagnosed each year. It ranks 7^th^ amongst cancer deaths with a reported 500,000 deaths annually. In India, with 52,396 new patients diagnosed and 46,504 deaths each year, EC ranks 8^th^ amongst cancer incidence and 7^th^ common cause of cancer death(1). There are two major histological types, squamous cell carcinoma and adenocarcinoma and each may represent two distinct diseases with different pathogenesis, epidemiology, prognosis, and tumor biology, including the pattern of lymph node metastasis. In East Asia, squamous cell carcinoma (ESCC) is the most common type, whereas adenocarcinoma is predominant in Western countries(2).

Surgery has been regarded as mainstay of cure for esophageal cancer in the past. Advances in surgery, radiotherapy (RT), and chemotherapy have established multimodality treatment options for EC. While neoadjuvant therapy followed by surgery is the treatment of choice for operable EC, definitive chemo-radiotherapy (dCRT) is the preferred treatment for inoperable patients, cases with unresectable EC, or patients who decline surgery.

The RTOG 85-01 trial established the role of definitive chemo-radiotherapy (dCRT) as standard for non-operative therapy of localized EC, reporting a median survival of 12.5 months for those receiving chemo-radiotherapy versus 8.9 months with radiotherapy alone(3). Randomized studies have subsequently confirmed these findings with survival rates of 20-25% at 5 years with dCRT, although recent studies utilizing modern radiation techniques have shown higher survival rates(4). High rates of locoregional failure (41–50%) after dCRT are common while using conventional radiation doses of 50.4Gy for EC, with the vast majority of failures involving the primary tumour (86–90%)(5).

To tackle the high locoregional failures, additional strategies that have been studied include induction chemotherapy, higher radiation dose, integration of new generation chemotherapy agents and targeted therapy. However, many of those strategies have resulted in increased toxicity and treatment-related mortality and no improvement in locoregional control rates or survival. Two-dimensional (2D) treatment planning using conventional methods treated larger volumes, which increased the risk of serious pulmonary and esophageal toxicity. Studies using 3D conformal radiotherapy (3DCRT) or intensity-modulated radiotherapy (IMRT) have shown improvement in target conformity and reduced exposure of normal tissues (e.g. lungs and heart) compared with 2D radiotherapy (2DRT) (6).

Standard dose of 50.4Gy in treatment of EC is based on the INT 0123 trial which, attempting a dose escalation to 64.8Gy along with cisplatin and 5FU showed no benefit with higher toxicity and on-treatment deaths in the dose escalated arm (7). Local failure rates of around 45% in both the RTOG 8501 and INT 0123 trials leaves much room for improvement.

The objective of the current study is to report the long term follow-up, toxicity, and outcomes with dCRT in patients with localized ESCC diagnosed at our institute.

## MATERIALS AND METHODS

Between January 2000 and March 2012, 545 patients diagnosed with carcinoma post cricoid, upper cervical and thoracic oesophagus treated with radical intent combination strategies of dCRT were identified from the hospital medical records. Of this, 314 patients with ESCC, who had completed the planned treatment and had at least been seen at one follow-up post treatment were included in the analysis. Demographic, disease, treatment and outcomes data were extracted from patient case files and hospital medical records. Those patients whose information on disease status were not available were contacted telephonically or by using postal service. The study was approved by the Institutional review board.

An earlier study from our institution had reported outcomes in patients who were treated between 2011 and 2014(x). The present study includes, in addition to those treated before 2010 at our institution, few of these patients.

All patients underwent endoscopic evaluation and had a tissue diagnosis. Patients underwent computed tomography of chest and abdomen (and also the neck in patients with cervical and upper esophageal cancer). Evaluation with ^18^F–fluoro-deoxy-glucose positron-emission tomography (PETCT) scan was introduced after 2009. The disease was initially staged according to the 2009 TNM classification and stage grouping(9). Patients were reclassified according to the 8^th^ edition AJCC for the purpose of this analysis(10). Patients deemed inoperable (surgical or medical reasons, or patient refusal) underwent upfront RT with or without concurrent chemotherapy. Patients with borderline resectable and/or bulky disease received neoadjuvant chemotherapy followed by CRT or RT alone.

Radiotherapy was delivered in 2 phases; in phase I, primary disease with a 5cm longitudinal margin and involved nodes were treated to a dose of 39.6 Gy in 23 fractions. In case of cervical esophagus or upper thoracic disease, bilateral supraclavicular fossae were prophylactically treated, and celiac nodes were included in lower thoracic esophageal cancers. In Phase II, a dose of 19.8 – 24 Gy in 11-12 fractions was delivered. Neoadjuvant chemotherapy schedule given was either a doublet combination of Cisplatin and 5-Flurouracil or Paclitaxel and Carboplatin. Weekly Cisplatin or Paclitaxel / Carboplatin was given concurrently during radiation.

### Response to dCRT and follow-up

Evaluation of treatment response was carried out 2 months after completion of treatment. Response was assessed by upper GI endoscopy (UGIES), with biopsy from any suspicious lesion, if present and contrast enhanced CT scan of the thorax. Acute and late radiotherapy toxicity assessment was done according to RTOG criteria. Follow-up was done quarterly thereafter. On follow-up, a barium swallow study, UGIES or CT evaluation of the thorax/abdomen were carried out as clinically indicated. A local failure was recorded if there was a positive pathological diagnosis (on endoscopic biopsy) or evidence of recurrence shown on CT/ Ba swallow evaluation. Local relapses were defined as those occurring within the mediastinum or within irradiated portals. Patterns of recurrence were defined as locoregional, distant (metastatic) or both locoregional and distant, when both were diagnosed at the same time. The time of recurrence was taken as the date of the confirmatory investigation.

### Statistical Analysis

The reverse Kaplan–Meier method was used to estimate median follow-up time. Time-to-event outcomes were defined as the time from pathological diagnosis to the event. In the absence of an event, overall survival (OS), local and distant recurrence were censored on the date of the last follow-up. Progression free survival (PFS) was defined from the date of registration to date of any recurrence or last follow-up. OS was defined from date of registration to the date of death or date of last follow up. PFS and OS were calculated by Kaplan-Meier method of survival analysis and comparison between different subgroups was performed using the log rank test. Comparisons having a p value less than 0.1 on univariate analysis were selected for a multivariate Cox proportional hazards model. A 2-sided p value less than 0.05 was considered statistically significant. Data were analyzed using SPSS Version 22.

## RESULTS

### Patient characteristics

The median age at presentation was 56 years (range – 22 to 79 years) with a male: female ratio of 1.7:1 (63% males). Dysphagia (64 %) and weight loss (21%) were the most common presenting symptoms. The median duration of symptoms was 3 months (ranging from 1 to 9 months). Tobacco chewing was the most commonly associated lifestyle habit (38%) followed by tobacco and alcohol (14%) while 20% patients had no habits. The median Karnofsky Performance Status (KPS) at presentation was 70 (range: 60-90). Patients’ characteristics are presented in table 1.

**Table 1.** Patient characteristics (n = 314)

| **Table 1. Patient characteristics (n = 314)** |  |
| --- | --- |
| **Age** (years) | Median – 56 (Range 22-79) |
| **Gender**<br>Male<br>Female | 197 (63%)<br>117 (37%) |
| **Tobacco chewers**<br>Present<br>Absent | 242 (77%)<br>72 (23%) |
| **Symptoms**<br>Dysphagia<br>Weight loss<br>Hoarseness | 201 (64%)<br>66 (21%)<br>09 (3%) |
| **Dysphagia to**<br>Solids<br>Liquids | 207 (66%)<br>107 (34%) |
| **Symptom duration**(months) | Median 2 (range 1-9) |
| **Karnofsky Performance score** | Median 70 (range 60-90) |

### Disease characteristics (Table 2)

Apart from 16% patients who had disease in the cervical esophagus, all other patients had disease in the distal esophagus with the thoracic esophagus being involved in 43% of patients. Most patients had T3 disease (58%) with 43% having N1 nodal disease at presentation.

**Table 2.** Disease and treatment characteristics (n = 314)

| **Table 2. Disease and treatment characteristics (n = 314)** |  |
| --- | --- |
| **Location** |  |
| Cervical | 50 (16%) |
| Upper Thoracic | 105 (33%) |
| Middle Thoracic | 133 (43%) |
| Lower Thoracic | 26 (8%) |
| **Bronchoscopy** |  |
| Abnormal | 51 (16%) |
| Normal | 263 (84%) |
| **Length of involvement** |  |
| < 7cm | 255 (81%) |
| >7cm | 59 (19%) |
| **T Stage** |  |
| T1 / T2 | 28 (9%) |
| T3 / T4 | 286 (91%) |
| **N Stage** |  |
| N0 | 147 (47%) |
| N+ | 167 (53%) |
| **Neoadjuvant chemotherapy** |  |
| Yes | 111 (35%) |
| No | 203 (65%) |
| **RT dose** |  |
| < 63 Gy | 166 (53%) |
| > 63 Gy | 148 (47%) |
| **Concurrent chemotherapy** |  |
| Yes | 178 (57%) |
| No | 136 (43%) |
| **Radiotherapy duration** | Average - 52 days (IQR 48 – 52 days) |

### Treatment details

Radiotherapy was delivered using a combination of conventional and conformal techniques in 198 patients (63%). About 35% patients received neoadjuvant chemotherapy prior to radical radiation therapy; Platinum and taxane combination was the most common (80%) regimen administered and 80% received two or more cycles. About 57% patients received concurrent chemotherapy during radiation. Concurrent chemotherapy was delivered as a weekly regimen (64%) and most patients received single agent platinum (46%) as a concurrent agent and while 16% patients received platinum/5FU combination regimen. About two thirds of patients received at least 4 cycles of concurrent chemotherapy.

Median dose of radiation delivered was 60 Gy (range 30.6 Gy – 70 Gy). Median overall treatment time of radiotherapy was 52 days (range 6 – 90 days). Radiotherapy techniques were conventional in 102(32%) patients, 3DCRT in 62(20%) and a combination of conventional and 3D-CRT in 143(45%). IMRT was used in only 8(3%) of the patients.

### Treatment toxicity

There were 242 patients (77%) who completed the treatment without a break and a further 12 patients (4.5%) who had a break due to machine issues but completed their planned treatment course. Thirty patients (10%) required hospitalization either due to electrolyte imbalance or febrile neutropenia; seven patients defaulted and did not complete treatment. Two patients (0.6%) died during treatment; one because of haematemesis leading to aspiration pneumonia and second due to very poor general condition and progressive dysphagia. During treatment, 112 patients (36%) required a naso-gastric tube placement; 5 patients had a percutaneous endoscopic gastrostomy performed and 3 required parenteral nutritional support.

Acute reactions on treatment were graded using the RTOG acute toxicity criteria. Most of the patients experienced only grade 1 or 2 (77%) with only 6 patients (2%) developing grade 3 dermatitis. Grade I mucosal reactions were seen in 94 (30%) and Grade II in 129 (41%) patients while grade 3 mucosal reactions were seen in 2 (0.6%) patients. With regards to hematological toxicity, grade 3 or above anemia, neutropenia and thrombocytopenia were seen in 2%, 4% and 1% respectively. The toxicity rates (acute and late) were not significantly different between RT doses (< 63 Gy vs ≥ 63 Gy). There was no dysphagia in 97 patients (31%) while 7% patients had chronic dysphagia of grade 2 or above. Although esophageal stenosis was seen in 55 patients, half of them were asymptomatic and did not require any chronic feeding procedure.

### Patterns of failure and Survival analysis

At 1^st^ follow-up, 175 patients (56%) had complete response, 114 (36%) had partial response, 15 (5%) had progressive disease and 6 (2%) had stable disease while response assessment was not available for 4 (1%) patients.

Median follow up for the entire cohort was 56 months. At the time of analysis, 77 (24.5%) patients were alive without disease. Ninety-two patients (29%) had died due to disease and 3 patients (1%) of other causes. The median OS was not reached for the entire cohort; however, 58 patients who had disease at last follow up and had not come for follow up for more than 12 months were presumed and coded dead at their last follow-up date. The median PFS was 28 months; 1-, 2- and 3-yr PFS were 66%, 52% and 46% respectively (Figure 1). Median OS was then found to be 28 months (Figure 2). The 1-, 2- and 3-yr OS were 80%, 67% and 62% respectively. A total of 152 patients (48%) had failed either locally or loco-regionally. 7 patients developed distant metastases; one patient developed a second primary.

**Figure 1a.**
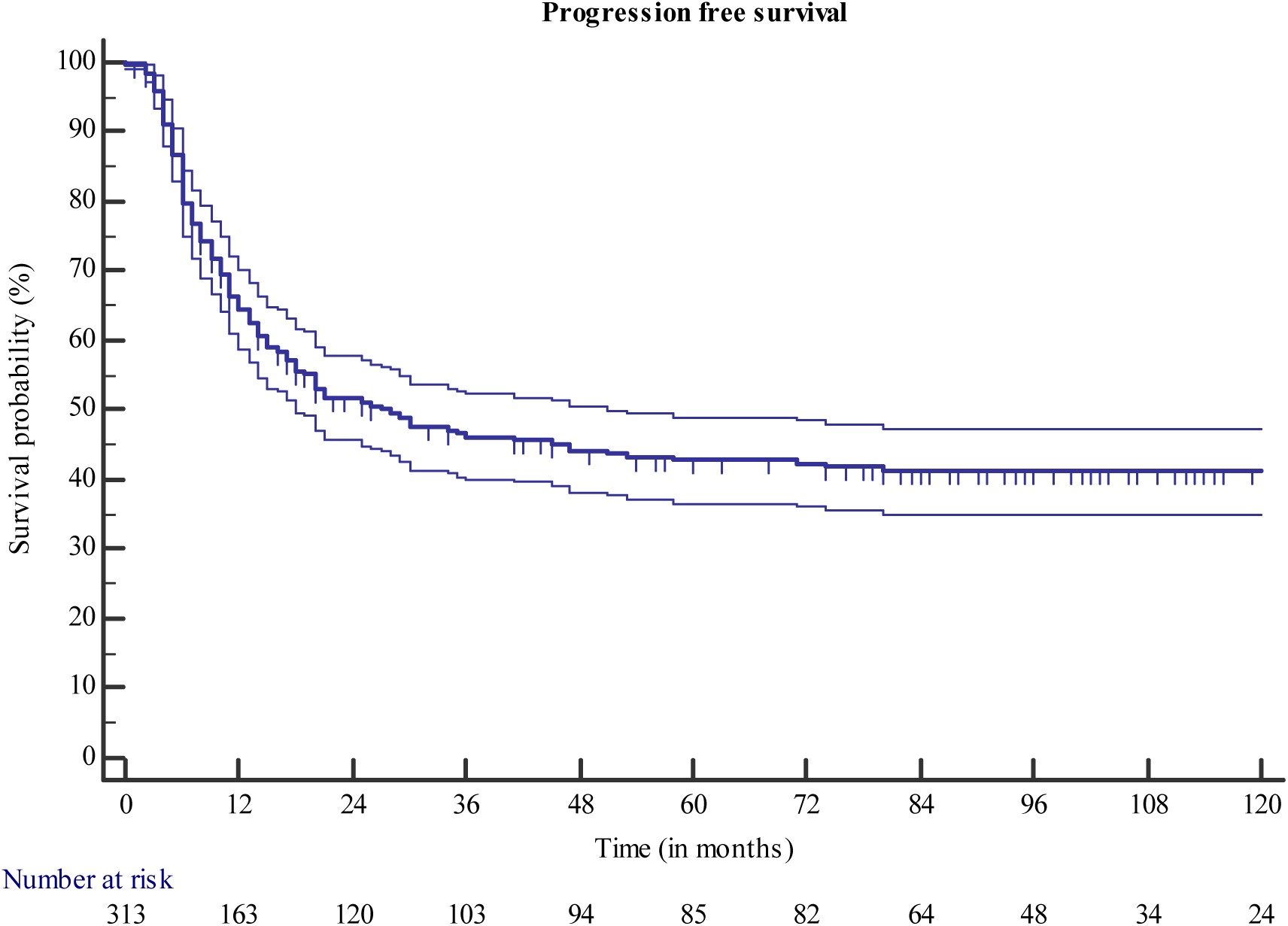
Kaplan–Meier’s curves for progression free survival (with 95% CI)

**Figure 1b.**
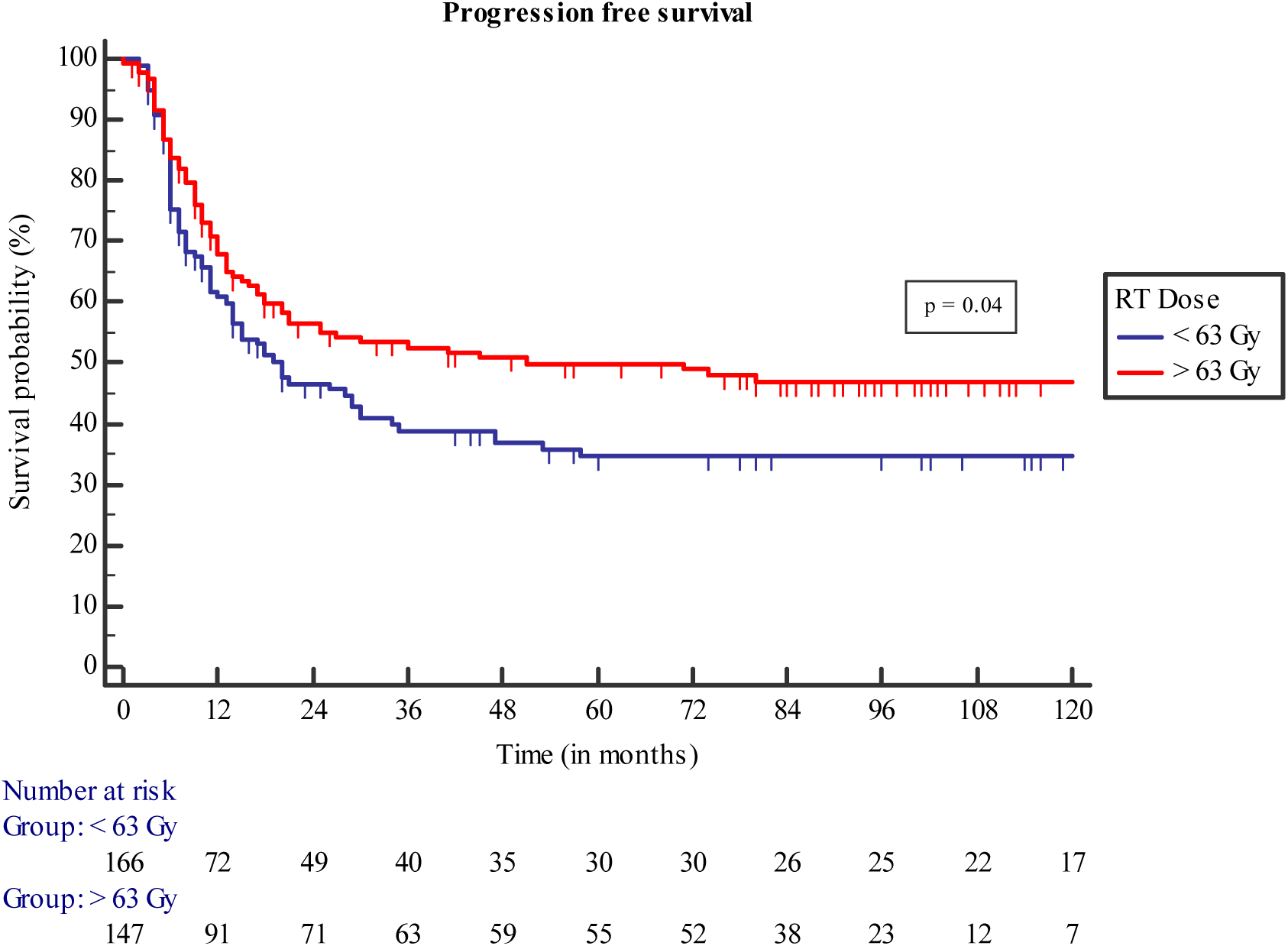
Kaplan–Meier’s curves for progression free survival of patients receiving different RT doses.

**Figure 2a.**
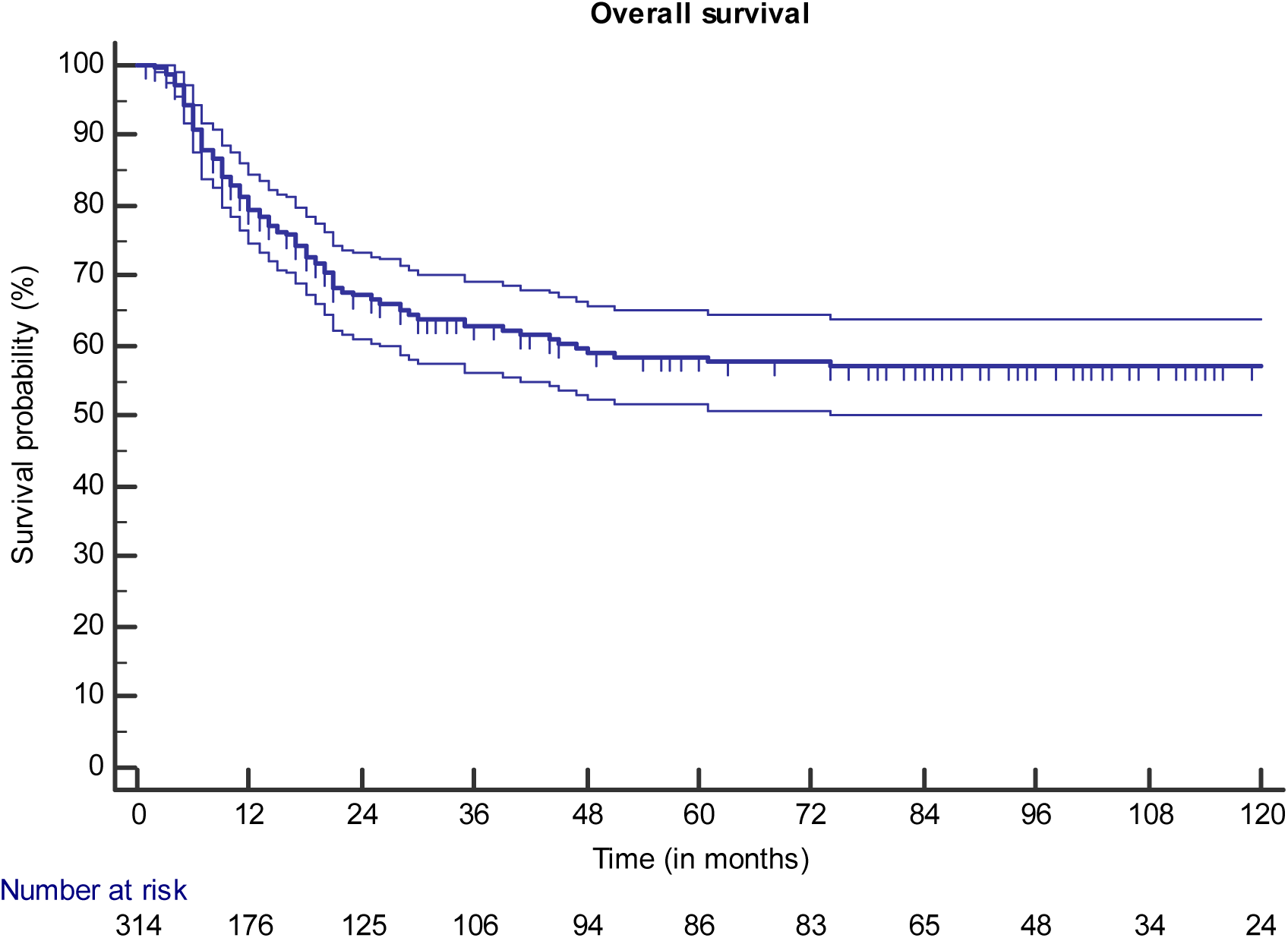
Kaplan–Meier’s curves for overall survival with 95% CI.

**Figure 2b.**
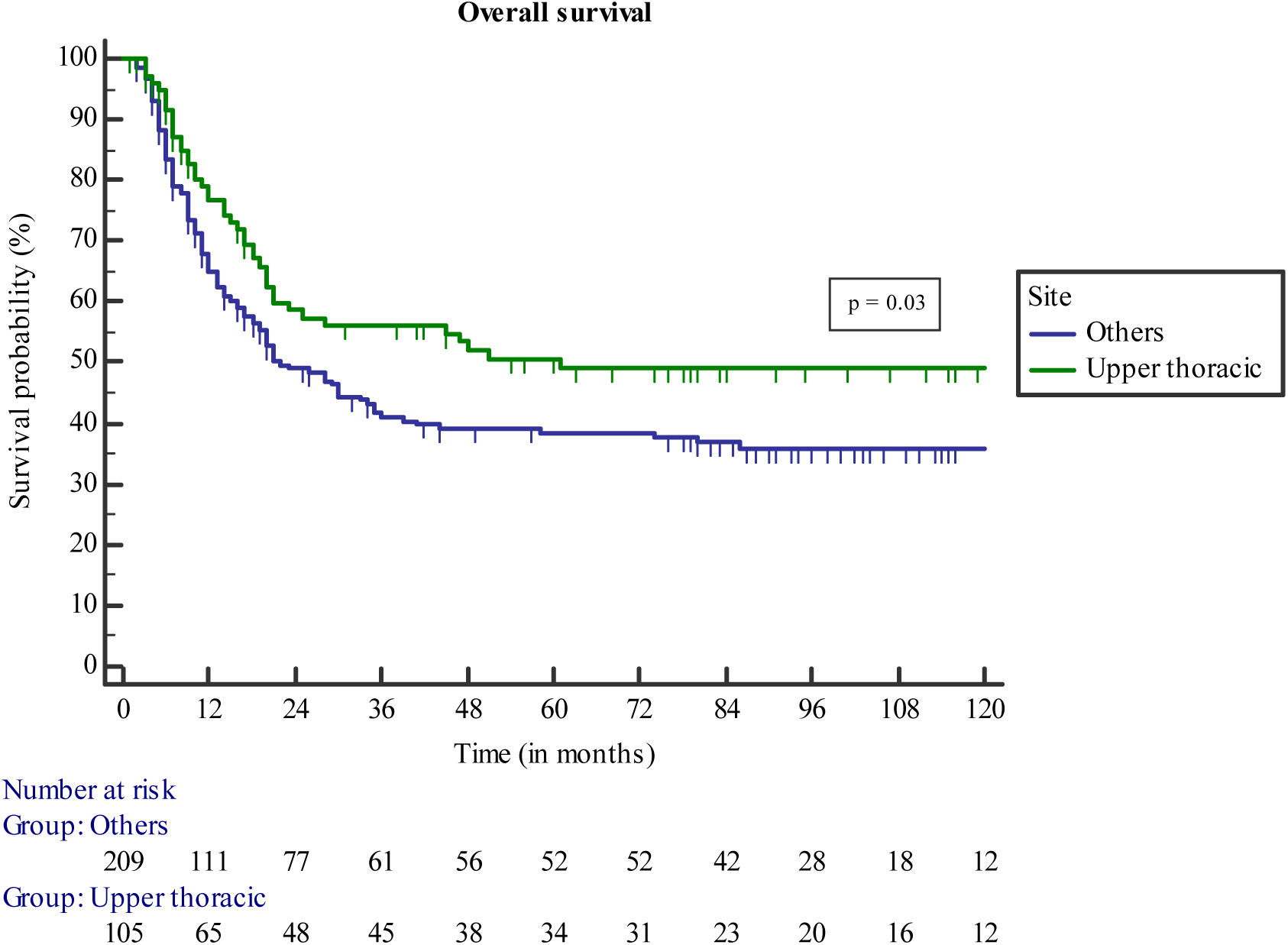
Kaplan–Meier’s curves for overall survival of patients according to primary site.

**Figure 2c.**
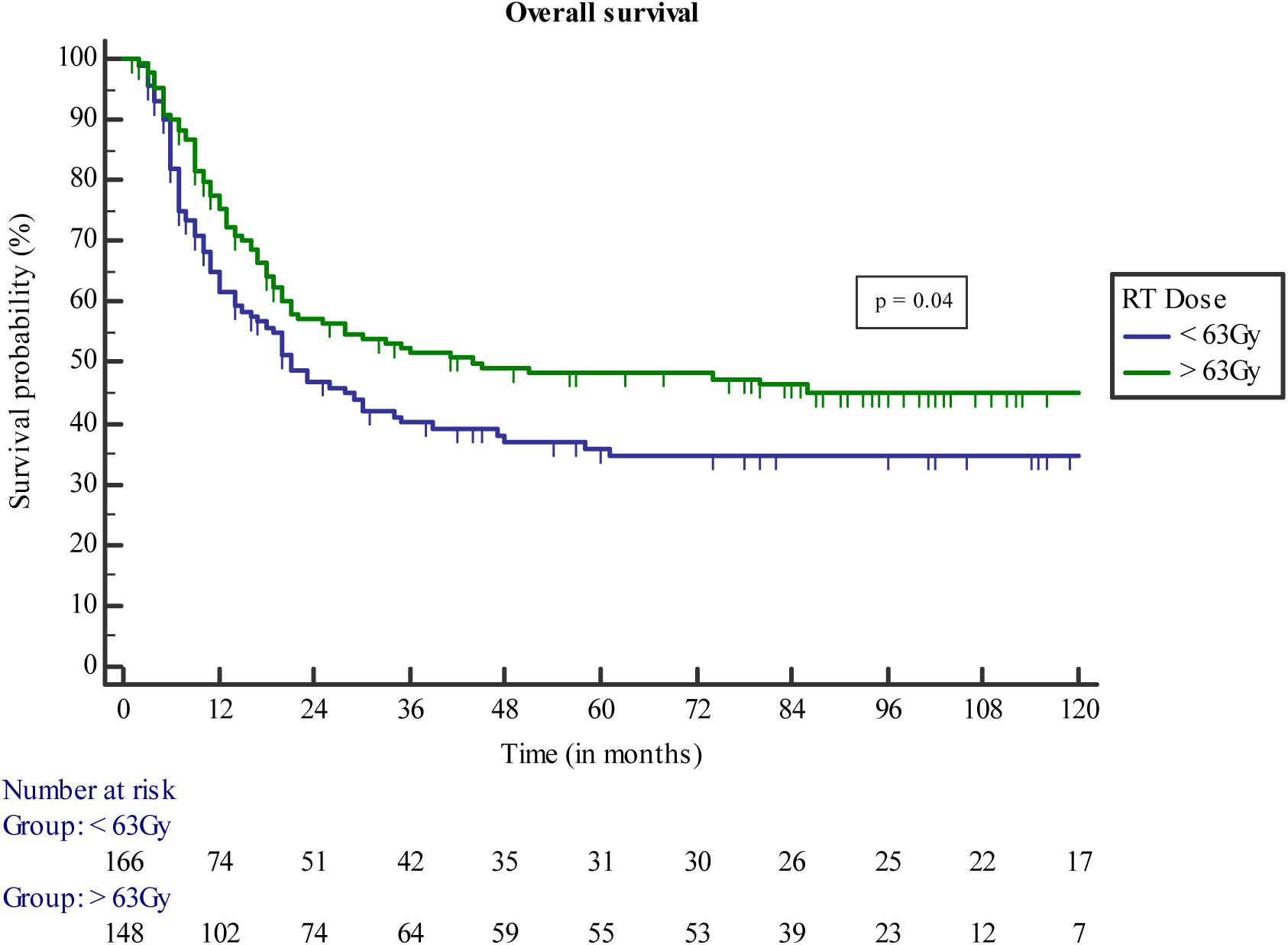
Kaplan–Meier’s curves for overall survival of patients receiving different RT doses.

On univariate analysis (Table 3), tobacco chewing, grade 2 dysphagia and above, T stage, nodal involvement, total dose of RT ≥ 63 Gy and concurrent chemotherapy were significant for PFS. For OS, additionally, a location of upper thoracic esophagus was significant. However, on multivariate analysis (Table 4), only T stage, nodal involvement and higher RT dose were significant for both OS and PFS.

**Table 3.**
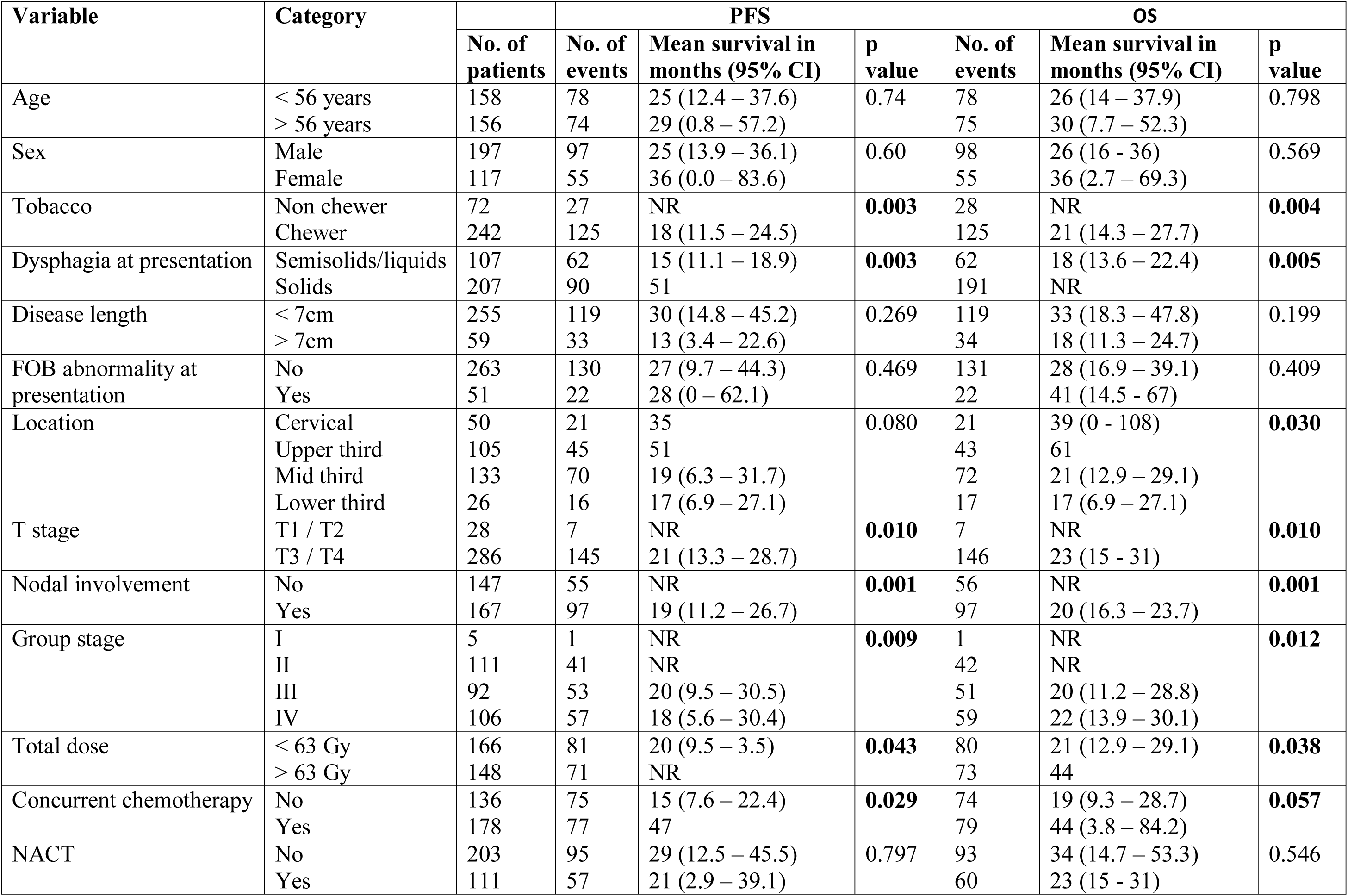
Results of the univariate analysis.

**Table 4.** Multivariate analysis.

| **Table 4. Multivariate analysis** |  |  |  |  |  |
| --- | --- | --- | --- | --- | --- |
| Variable | Category | PFS |  | OS |  |
|  |  | Mean survival in months (95% CI) | p value | HR (95% CI) | p value |
| Location | Others vs Upper esophagus | 1.37 (0.95 – 1.97) | 0.09 | 1.51 (1.04 – 2.18) | **0.03** |
| T stage | T1 / T2 vs T3 / T4 | 0.42 (0.19 - 0.91) | **0.02** | 0.41 (0.19 – 0.88) | **0.02** |
| Nodal involvement | No vs Yes | 0.63 (0.45 – 0.88) | **0.007** | 0.65 (0.47 – 0.91) | **0.01** |
| Total dose | < 63 Gy vs > 63 Gy | 0.72 (0.52 – 0.99) | **0.04** | 0.72 (0.52 - 1) | **0.03** |
| Concurrent chemotherapy | No vs Yes | 1.24 (0.89 – 1.73) | 0.20 | 1.16 (0.84 – 1.62) | 0.36 |

## DISCUSSION

The current study investigated the relationship between various disease and treatment parameters and their correlation with survival among patients with inoperable, non-metastatic EC who underwent definitive RT with chemotherapy. The analysis of this retrospective dataset indicates that a total RT dose of ≥63□Gy to the primary tumour was associated with significantly better PFS and OS than a lower dose. This difference persisted even after accounting for patients receiving concurrent chemotherapy (p = 0.023).

Management of patients with unresectable or inoperable esophageal cancer is challenging and requires a multimodality approach(11–13). This is a heterogeneous group that includes patients with potentially resectable (T4a, and unresectable (T4b) primary disease, poor surgical candidates, and those who refuse surgery(13,14). Most of these patients in addition to having loco-regionally advanced disease are also nutritionally compromised and suffer from significant comorbidities, making it difficult to successfully complete planned treatment(14).

The CROSS trial established triple modality using neoadjuvant chemo-radiation (nCRT) followed by surgery as the standard of care in operable EC. This Dutch multi-center randomized controlled trial compared surgery with or without nCRT in 366 patients with EC. After nCRT, 92% of the patients underwent a R0 resection compared to 69% in the surgery alone group. Pathological complete response (pCR) was seen in 29% of the patients (23% in EAC and 49% in ESCC). Also, 5-year OS improved from 33 to 47% with no increased postoperative complications in the patients undergoing nCRT(15). A similar trial for patients with exclusively ESCC alone, conducted in China, the NEOCRTEC5010 trial, showed near identical pCR of 43% and median OS of 100 months(16). These impressive R0 resection, pCR and survival rates after nCRT highlight two important beliefs. One, given that roughly 50% patients with ESCC had no disease left after nCRT, is there a need for surgery at all? Two, since only 40-41.4 Gy was used in the neoadjuvant strategy, escalating to higher doses of RT may consolidate the loco-regional disease control and obviate the need for surgery.

Two large randomized trials challenged the role of surgery in potentially resectable esophageal cancer. Stahl et al. randomized 172 patients with locally advanced ESCC to nCRT followed by surgery and dCRT. Although the PFS was better in the surgical group (64.3 vs. 40.7%; HR 2.1, p = 0.003), treatment-related mortality was higher (12.8 vs. 3.5%; p = 0.03) and 3-yr OS was similar in both groups(17). Bedenne et al. in a similar design, randomized 259 patients with locally advanced EC; although local recurrence rate after 2 years was higher in the patients undergoing dCRT (HR 1.63, p = 0.03), mortality in the first 3 months postoperatively was higher in the surgery arm (HR 1.63, p = 0.002)(18). A recent meta-analysis showed that dCRT and surgery are equally effective as initial treatments for resectable esophageal cancer(19). Transhiatal esophagectomy poses significant risks, including tracheal and pulmonary injury, anastomotic leak, vagus nerve injury, infection, and death(20). Currently, based on the finding of a similar outcome in survival in the randomized trials, the approach of dCRT is a reasonable choice especially in patients with ESCC. Recently, Dutch investigators attempting to identify the accuracy of detecting residual disease after nCRT showed feasibility with different diagnostic approaches and are conducting a phase 3 randomised controlled trial to assess active surveillance with this combination of diagnostic modalities. This approach, if successful, would help identify patients who could potentially avoid surgery(21).

Based on the INT 0123 trial, the NCCN recommends a radiation dose of 50 or 50.4 Gy for dCRT. This trial compared high and standard doses (64.8□Gy vs. 50.4□Gy, for locally advanced EC, and was prematurely closed because of multiple deaths in the high-dose group and no differences in survival or local control. However, 7 of the 11 treatment-related deaths in the high-dose group occurred before the radiation dose reached 50.4□Gy, and the high-dose group also received a significantly lower dose of fluorouracil, which might have affected outcomes in that arm. Moreover, the investigators used conventional techniques to deliver RT(7). Modern RT techniques, such as 3DCRT and IMRT or volumetric modulated arc therapy (VMAT) have shown to widen the therapeutic ratio effectively(6).

Various other studies have evaluated RT dose escalation in EC with equivocal results. In an analysis of the National Cancer Database in the US, higher RT doses (51–54, 55–60, or >60 Gy) did not improve OS relative to a dose of 50–50.4 Gy(22). Chen et al. performed a population-based propensity score-matched study using Taiwanese registry data for patients with EC who underwent IMRT or 3DCRT and reported that a higher RT dose of ≥60 Gy provides better 5-year OS than 50–50.4 Gy (22% vs. 14%, p < 0.05)(23). Another group studying the same cancer registry, analyzed over 2000 patients with ESCC receiving IMRT based concurrent chemo-radiation. On multivariate analysis, they found that a dose of more than 60 Gy was a significant independent prognostic factor for overall survival(24). In a systematic review, Song et al. reported that dCRT to a dose of >60 Gy), provided better tumour response, locoregional control, and OS(25). These diagonally opposite conclusions between the orient and occident populations could be attributed to the difference in the predominant histologies of EC. For example, most patients in the NCDB registry had adenocarcinoma histology (58%) and higher dose was associated with better overall survival on univariate but not multivariate analyses. A further disparity could lie between the ESCC histology that is encountered in the west and east. The Cancer Genome Atlas Research Network in their seminal work, performed a comprehensive molecular analysis and divided ESCCs into three subtypes: ESCC1, ESCC2 and ESCC3. These subtypes showed trends for geographic associations with tumours from Asian population studied, tended to be ESCC1 and European and American patients’ tumours tended to be ESCC3(4,26). This geographic difference could be the key to the difference in the clinical results between the east and west. Our results also support this association between improved OS and higher RT doses (≥63 Gy), relative to doses of <63 Gy.

The present study also demonstrates that unlike RTOG 94-05, higher RT dose was well tolerated as reflected by the compliance and toxicities. Majority (77%) of the patients completed their planned treatment without any interruption, 30 (10%) patients were hospitalized during course of treatment but completed the planned treatment after recovering from acute illnesses. This higher compliance to treatment could possibly be due to the smaller target volumes unlike the earlier RTOG trials where prophylactic nodal irradiation was practiced(3,7), omission of infusional 5-FU and use of single agent platinum in majority of patients (103/149 - 70%) in the concurrent setting leading to better tolerability(27). With paclitaxel and carboplatin concurrent regimen, we had earlier reported a clinically significant incidence in grade 3 toxicity of 56% with 60% patients developing chronic toxicity of any grade(8). Although there are no published head to head comparison between 5-FU based and paclitaxel/carboplatin regimens, the latter is associated with lesser hematological toxicities and better tolerated(28). In the present report, grade 3 non-hematological or hematological toxicities were an infrequent occurrence; however, complete data on chronic toxicity was unavailable for a detailed analysis. Advances in delivery techniques have also led to further decrease in treatment related toxicities and reduced potential for marginal misses thereby improving both compliance and local control(6,29). Concurrent chemotherapy had a significant impact on DFS and OS on univariate analysis, but not on multivariate analysis, possible because of multi-collinearity with other unadjusted correlated variables.

Clinical staging is never as accurate as surgical staging (30) but continues to remain an important prognostic factor. In the present study PFS and OS were influenced significantly on univariate analysis by T (p<0.005) and N stage (<0.05) which retained significance on multivariate analysis.

The limitations of our study include its retrospective nature, lack of detailed toxicity and quality of life data. Also, the study analyzed only those patients for whom all treatment related information was available, the associated biases, and the fact that techniques for disease staging and treatment improved considerably over the long-time span of the study. In contrast the study has several strengths, in that it represents the largest series of consecutive patients treated over 12 years by an experienced multidisciplinary team of oncologists, out of a strict protocol setting.

In conclusion, this is one of the largest series examining the outcomes and identifying prognostic factors in patients with ESCC treated with definitive non-surgical methods in a non-trial, service setting. The study’s results suggest that higher RT doses (≥63 Gy), are feasible in this patient group and importantly, the higher RT doses were associated with significantly better PFS and OS than for lower doses.

## Data Availability

All relevant are available for review

